# A prebiotic diet intervention can restore faecal short chain fatty acids in Parkinson’s Disease yet fails to restore the gut microbiome homeostasis

**DOI:** 10.1101/2024.09.09.24313184

**Authors:** Janis Rebecca Bedarf, Stefano Romano, Silke Sophie Heinzmann, Anthony Duncan, Maria H. Traka, Duncan Ng, Daniella Segovia-Lizano, Marie-Christine Simon, Arjan Narbad, Ullrich Wüllner, Falk Hildebrand

## Abstract

Despite extensive research, current treatment of Parkinson’s Disease (PD) remains symptomatic and disease modifying approaches are urgently required. A promising approach is to target the gut-brain-axis by modifying the intestinal microbiota and the herein produced metabolites.

We decided to test this approach by modifying key metabolites of bacterial intestinal fermentation: short chain fatty acids (SCFA), known to be decreased in PD patients. A prospective, controlled pilot study was conducted in 11 couples, with one PD patient and healthy spouse as control (CO) each. Participants followed a 4-week diet rich in dietary fibre in addition to the intake of a prebiotic sirup (Lactulose). Metagenomes and metabolites of the gut microbiota, urinary metabolites and clinical characteristics were assessed.

The short-term dietary intervention significantly augmented gastrointestinal SCFA production, likely associated with increased *Bifidobacteria spp*. PD associated gastrointestinal symptoms improved with increasing SCFA levels. The pre-existing bacterial dysbiosis associated with PD, such as depletion of genera *Blautia*, *Dorea*, and *Erysipelatoclostridium* in PD, persisted within the study period. Some pathobionts, i.e. *Klebsiella,* were reduced after the intervention. Bacterial metabolite composition (both faecal and urine metabolomes) shifted towards the composition of the healthy control in PD after the intervention. Among these brain-relevant gut metabolic functions improved in PD patients, such as S-Adenosyl methionine (SAM), 3,4-Dihydroxyphenylacetic acid (DOPAC), Glutathione (GSH), Tryptophan and inositol related changes, involved in neuroprotective and antioxidant pathways.

Despite the small cohort size and short-term study period a minor dietary intervention was sufficient to improve gastrointestinal symptoms in PD and altered metabolic parameters in a presumed neuroprotective manner, warranting further investigation in larger cohorts.

## Introduction

The human gut microbiota form a complex community with high taxonomic diversity and complex metabolic activity. The bi-directional microbiota-related gut-brain signalling has emerged as an important factor in human brain (patho-)physiology^1^. The gut microbiome is involved in immune homeostasis and might have a role in the development or progression of neuro-psychiatric diseases, including multiple sclerosis, Alzheimer’s and Parkinson’s disease (PD)^2,3^ . The early involvement of the gastro-intestinal tract in PD – sometimes preceding motor symptoms for years – has been linked to the intestinal dysbiosis in multiple independent cohorts (reviewed in^4,5^). Taxonomic alterations hinted at impairments of the intestinal barrier and altered immune function as well at metabolic changes^6^ but it remains unclear, whether these changes are the causes or consequences of disease.

Among the variety of metabolites that have been shown to beneficially impact both intestinal barrier and blood brain barrier (BBB)-integrity^7–9^, short chain fatty acids (SCFA, represented mainly by butyrate, propionate, and acetate, and fermented from dietary fibres^10^) seem to play a pivotal role. For example, SCFA’s can directly influence the colonic epithelium, act as major energy substrates for colonocytes and promote tight junction proteins and neuronal activation through stimulation of G protein– coupled receptors^11,12^. Administration of butyrate in transgenic animal models of PD improved motor deficits, reduced inflammation and alleviated dopamine deficiency^13,14^ and together with a high-fibre diet, reduced markers of cerebral inflammation in aged mice^15,16^. SCFAs may thus present a link between gut microbiota dysbiosis and neurodegenerative alterations^12^. In line with a concept of anti-inflammatory and neuroprotective effects of SCFA, studies in human PD found depleted faecal SCFA concentrations and reduced abundance of SCFA producing bacteria compared with healthy controls^17,18^. Moreover, an integrated study examining both faecal and plasma SCFA levels, suggested that SCFAs may even reflect disease severity and could thus serve as a surrogate marker for PD^19^. In concordance with this data, a high fibre ‘mediterranean’ diet was reported to decrease the risk for PD^20,21^, in comparison to a low fibre, high caloric ‘western’ diet^22,23^.

Altering the microbiome composition and metabolism through dietary interventions might thus evolve as a therapeutic option to protect against or modulate the course of PD and other neurodegenerative diseases^24,25^. We here investigated whether a dietary intervention designed to prebiotically enrich colonic SCFA production would alter faecal SCFA concentrations and ameliorate gut microbiome dysbiosis as well as gastrointestinal symptoms in PD.

## Results

### Participants and clinical measures

10 of 11 enrolled couples (each consisting of 1 PD and 1 CO) completed the study (clinical characteristics and dietary baseline information in Suppl. Table 1) and performed the dietary intervention for 4 weeks (Fig. 1a). One couple dropped out of the study before the second visit due to non-study related reasons. PD patients were mild to moderately advanced (Hoehn and Yahr 1-2).

**Figure 1.**
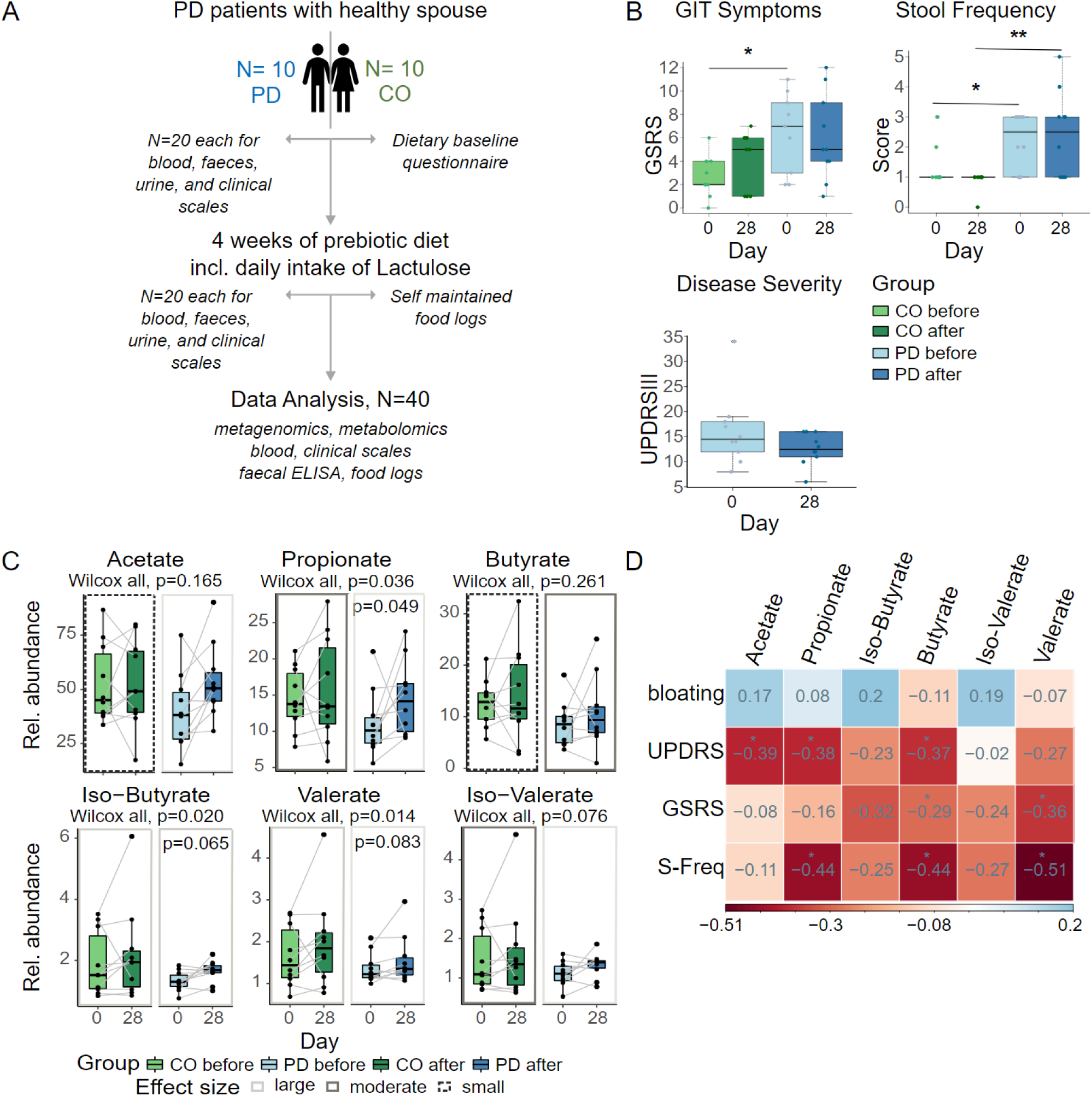
SCFA and clinical scales. A, study design; B, clinical measures, disease severity measured with UPDRSIII, gastrointestinal symptom scale (GSRS) with improvement in PD after prebiotics, and stool frequency score before and after prebiotics. * = p<0.05, ** = p<0.01; C, targeted SCFA measurements in faecal samples show increasing SCFA concentrations in both groups after prebiotics (Wilcoxon signed-rank test). Coloured borders indicate effect sizes; D partial correlation of SCFA concentrations with clinical measure shows a sign. inverse correlation of Acetate, Propionate, and Butyrate with disease severity measured with UPDRSIII, while stool frequency was inversely correlated to Propionate, Butyrate, and Valerate. GSRS shows a sign. negative correlation with Butyrate and Valerate, red= positive and blue= negative correlation, numbers indicate the correlation coefficient r.

**Suppl. Table 1.**
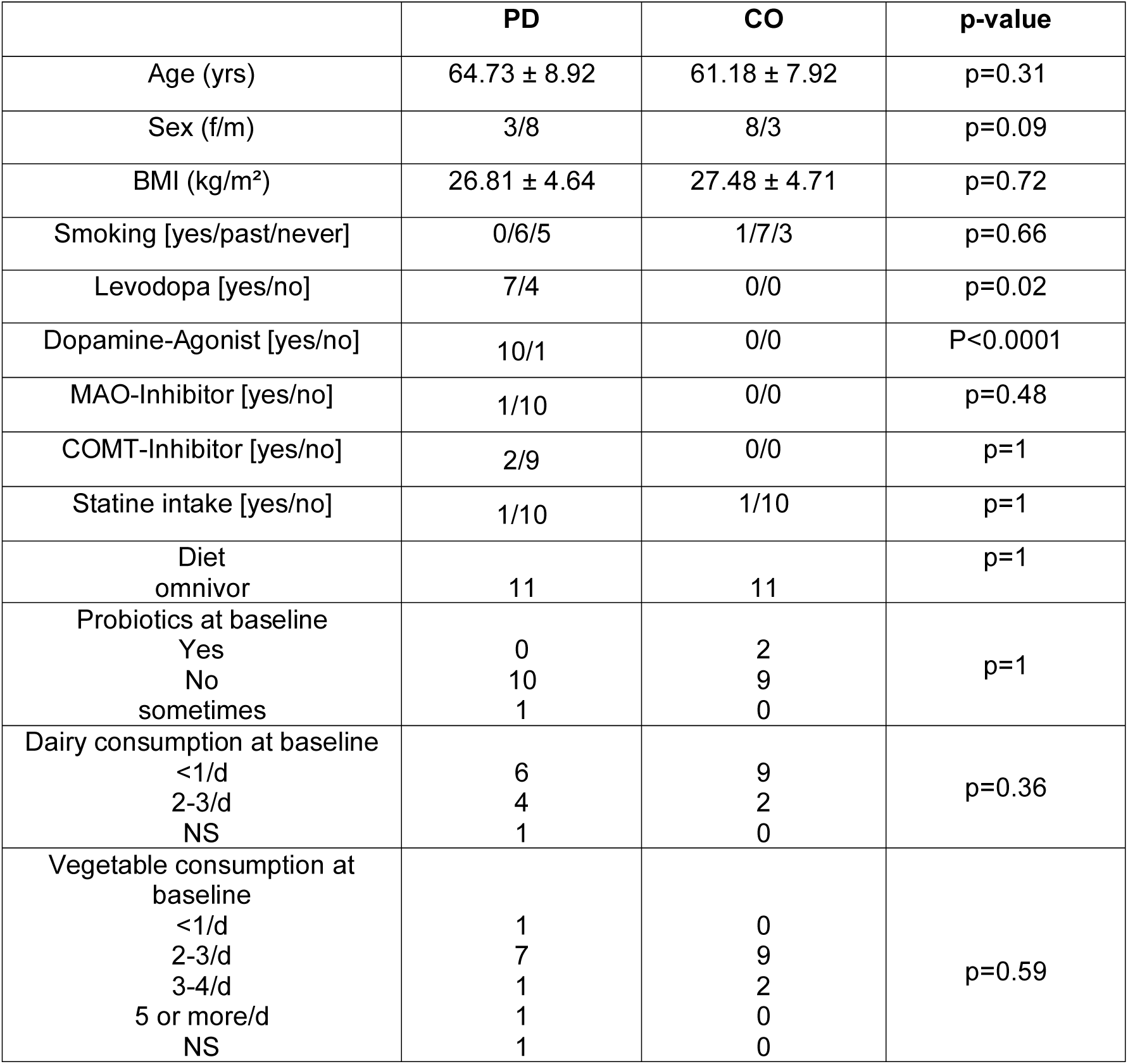
Clinical characteristics and dietary baseline information. Dietary baseline information was acquired by a questionnaire assessing the dietary habits of the participants before the intervention; Note that one couple dropped out of the study after baseline measurements due to non-study related reasons; BMI, body mass index; PD, Parkinson’s disease; CO, healthy control; Data is presented as mean ± standard deviation or absolute numbers of participants; NS, not specified.

|  | PD | CO | p-value |
| --- | --- | --- | --- |
| Age (yrs) | 64.73 ± 8.92 | 61.18 ± 7.92 | p=0.31 |
| Sex (f/m) | 3/8 | 8/3 | p=0.09 |
| BMI (kg/m<sup>2</sup>) | 26.81 ± 4.64 | 27.48 ± 4.71 | p=0.72 |
| Smoking [yes/past/never] | 0/6/5 | 1/7/3 | p=0.66 |
| Levodopa [yes/no] | 7/4 | 0/0 | p=0.02 |
| Dopamine-Agonist [yes/no] | 10/1 | 0/0 | P<0.0001 |
| MAO-Inhibitor [yes/no] | 1/10 | 0/0 | p=0.48 |
| COMT-Inhibitor [yes/no] | 2/9 | 0/0 | p=1 |
| Statine intake [yes/no] | 1/10 | 1/10 | p=1 |
| Diet omnivor | 11 | 11 | p=1 |
| Probiotics at baseline<br>Yes<br>No<br>sometimes | 0<br>10<br>1 | 2<br>9<br>0 | p=1 |
| Dairy consumption at baseline<br><1/d<br>2-3/d<br>NS | 6<br>4<br>1 | 9<br>2<br>0 | p=0.36 |
| Vegetable consumption at baseline<br><1/d<br>2-3/d<br>3-4/d<br>5 or more/d<br>NS | 1<br>7<br>1<br>1<br>1 | 0<br>9<br>2<br>0<br>0 | p=0.59 |

No serious adverse events occurred. Some participants (N=4 PD and N=4 CO) had tolerable bloating when starting the dietary intervention, which ceased during the study (self-reported); no diarrhea was reported. Safety parameters (sodium, potassium, C-reactive protein, and thyroid-stimulating hormone) were comparable between groups before prebiotics and there were no changes after prebiotics. All participants followed an omnivorous diet prior to the study with occasional intake of probiotics in 3 subjects (n=1 PD and n=2 CO, Suppl. Table 1).

To measure the participants’ compliance to the dietary recommendations, a food adherence score was calculated based on the recommended consumption of raw apples (see methods). This showed overall good adherence throughout the study cohort; four participants (n=2 CO and n=2 PD subjects, of whom 2 belong to one couple) had less compliance with our dietary intervention. (Suppl. Fig. 1a).

**Suppl. Figure 1.**
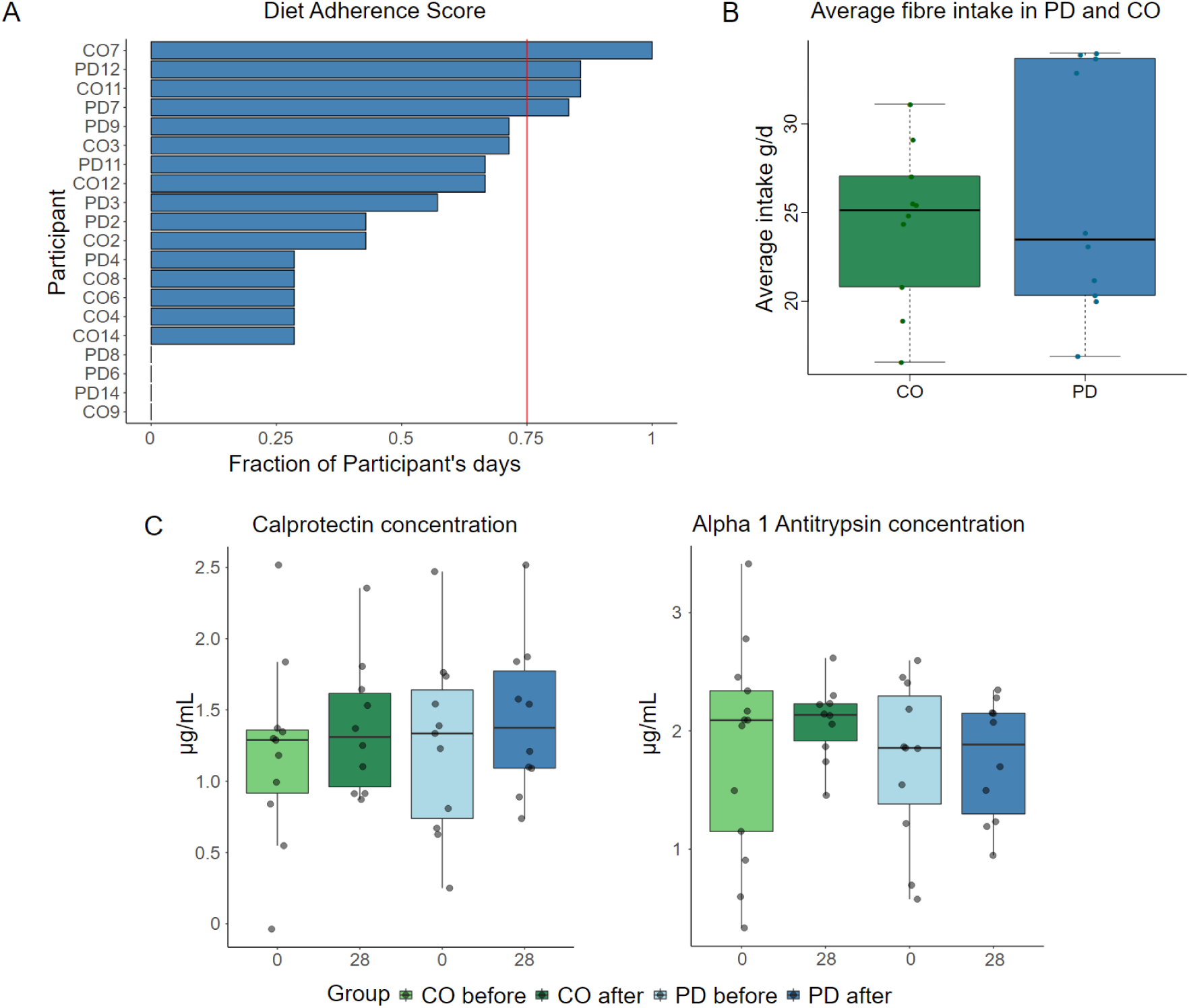
dietary compliance and gut integrity/inflammation markers. A, the dietary compliance score based on the consumption of apples shows n=4 individuals, who were less adherent to the recommended diet (data is shown as the fraction of participants’ diet days with an apple consumption below the mean apple consumption of all participants). A fraction of more than 0.75 days was considered as less adherent; B, average fibre intake was comparable in both study groups (Wilcoxon rank sum test, p>0.05); C, gastrointestinal markers of intestinal inflammation (faecal calprotectin, left) or intestinal permeability (faecal alpha-1-antitrypsin, right) were comparable between groups and did not change after prebiotics (Wilcoxon signed-rank test and Wilcoxon rank sum test, p>0.05).

The average fibre intake during the intervention was similar in both CO and PD (median 25.1 g/d and 23.5 g/d, respectively, Suppl. Fig. 1b). Although this is below the recommended daily intake of 30 g/d in Germany, it is still above average reported levels in Germany of 23 g/d and 20 g/d for males and females, respectively^26^.

UPDRS_III_, as a measure of the PD disease severity, was slightly reduced after the prebiotic intervention (Wilcoxon signed-rank test, p>0.05, Fig. 1b). While gastrointestinal symptoms (sum score of the modified GSRS) were significantly higher in PD patients compared to CO before prebiotics, no significant difference could be observed after prebiotics (Wilcoxon rank sum test, p=0.025 and p=0.48, respectively, Fig. 1b); however, this could be related to increasing GIT symptoms in the CO, mostly related to constipation and bloating. Stool frequency (as a sub measure of the GSRS) was sign. differed between PD and CO and this did not change during diet intervention (Wilcoxon rank sum test, p=0.043 and p=0.007, respectively, Fig. 1b). Faecal markers of intestinal inflammation (Calprotectin) or intestinal permeability (Alpha-1-Antitrypsin, Suppl. Fig. 1c) were comparable between study groups and remained unchanged after prebiotics.

### The prebiotic diet increased faecal SCFA levels and improved gastro-intestinal functioning in PD

Faecal SCFA concentrations were lower in PD subjects at baseline (Wilcoxon rank sum test, p<0.05, q>0.1), note that baseline SCFA results were comparable between targeted SCFA measures (Fig. 1c) and SCFA based on metabolomics (Suppl. Fig. 2a). The short-term dietary intervention increased concentrations of most SCFAs (Fig. 1c), as the statistical power was likely limited by cohort size.

**Suppl. Figure 2.**
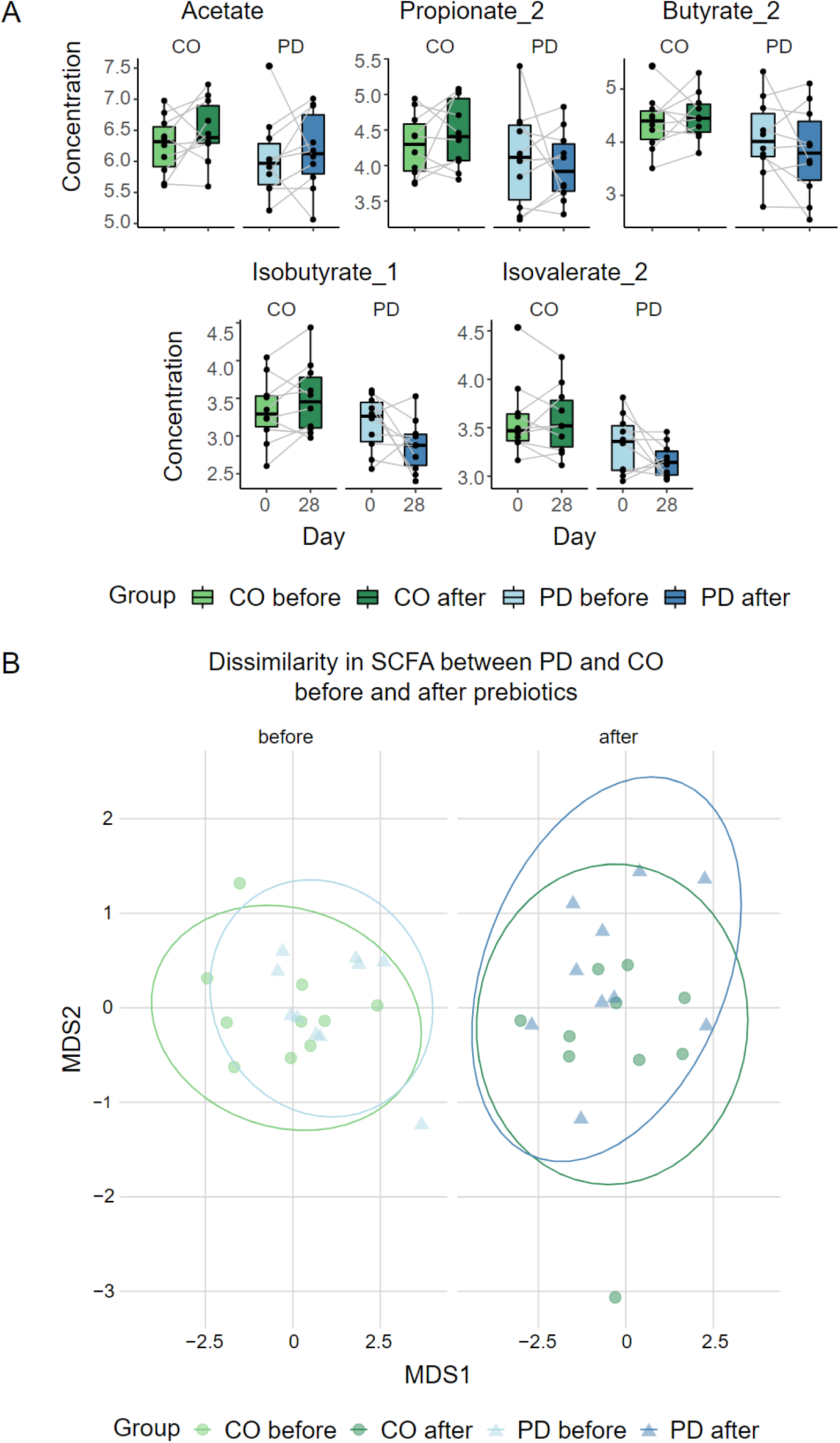
SCFA. A, relative abundance of SCFA based on metabolomics (log transformed data) shows comparable baseline conditions between PD and CO similar to targeted SCFA measures, B, dissimilarity of SCFA composition between PD and CO before and after prebiotics show that SCFA composition equalizes after prebiotics (perMANOVA, before R²=0.13, p=0.08, after R²=0.01, p=0.8, unconstrained dbRDA, Euclidean distance, targeted SCFA measures).

Considering effect sizes (borders in Fig. 1c), the increase in several SCFA concentrations was markedly greater in PD than in CO, possibly related to an increased Lactulose intake in the PD group.

Multivariate testing of the SCFA composition between PD and CO either before or after the diet intervention revealed fewer dissimilarities after prebiotics (perMANOVA before R²=0.13, p=0.08, and after R²=0.01, p=0.8, respectively, Suppl. Fig. 2b), indicating that the SCFA profile of PD patients adapted to a healthy state after prebiotics.

Stool frequency, modified GSRS, and PD disease severity were inversely correlated with several SCFA including butyrate, in PD patients (Fig. 1d), suggesting that increased SCFA concentrations were linked with better gastro-intestinal functioning and reduced disease symptoms.

### The prebiotic diet enhanced beneficial Bifidobacteria in both study groups yet failed to restore the dysbiosis associated with PD

All 40 faecal samples from 20 participants and two time points each, were sequenced using short read metagenomics. MGS (metagenomic species) dereplicated from MAGs (metagenomics assembled genomes) were used to obtain the taxonomic composition for all samples.

Species level alpha diversity was comparable between study groups, however, richness and diversity decreased after prebiotics (only sign. for CO, Wilcoxon signed-rank test, p=0.02, Suppl. Fig. 3a). The genera *Bifidobacteria, Bacteroides, Faecalibacterium, Blautia A,* and *Phocaeicola* dominated the overall taxonomic composition (Suppl. Fig. 3b).

**Suppl. Figure 3.**
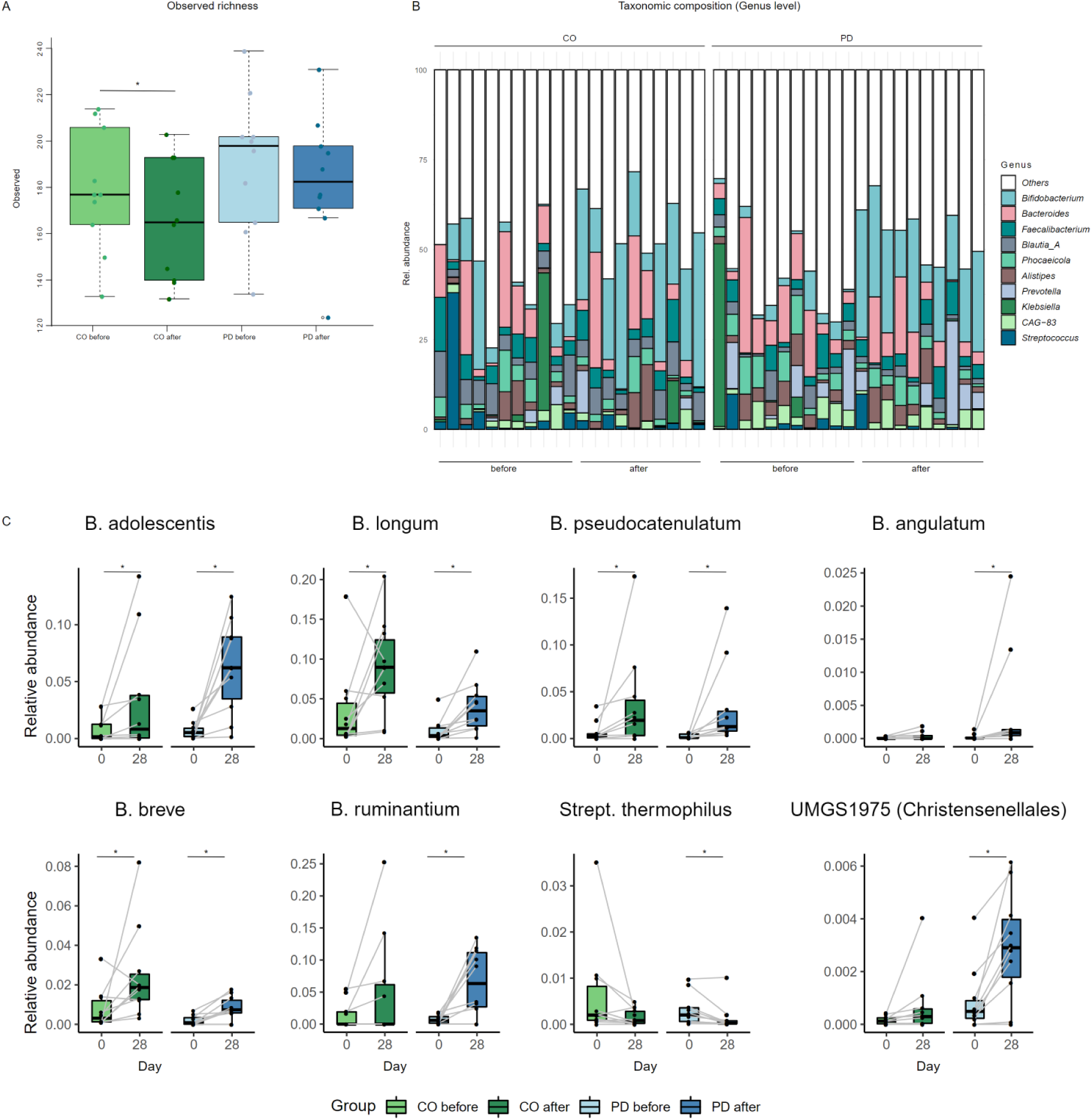
Richness and bifidogenic effect of the prebiotic intervention. A, Species level alpha diversity was not sign. different between PD and CO. However, within the CO group, species richness decreased after prebiotics (p=0.02, Wilcoxon signed-rank test); B, Composition of the top 10 Genera, separated for PD and CO; C, before vs. after comparisons (Wilcoxon signed-rank test revealed that several Bifidobacteria spp. increased after prebiotics, boxplots show relative abundance of taxa. n=6 *Bifidobacteria spp.* were enriched in PD after prebiotics, of which n=4 were also enriched in CO individuals; n=2 taxa were only sign. different in PD (Strept. Thermophilus, UMGS1975).

Most of the species level differences in faecal sample composition were driven by household (R² = 0.47, p<0.001, perMANOVA, dbRDA), i.e., most samples of the same household clustered together (Fig. 2a). Disease status (PD vs.CO) and diet intervention (before vs. after prebiotics) explained only 9.8% of total variance together (Fig. 2b and c). This “household-effect” was the prevalent source of variation between individual gut microbiomes at all taxonomic levels (R2 = 0.41 – 0.47) as well as functional potential and metabolomics profiles (R2 = 0.36 – 0.47, Suppl. Table 2).

**Figure 2.**
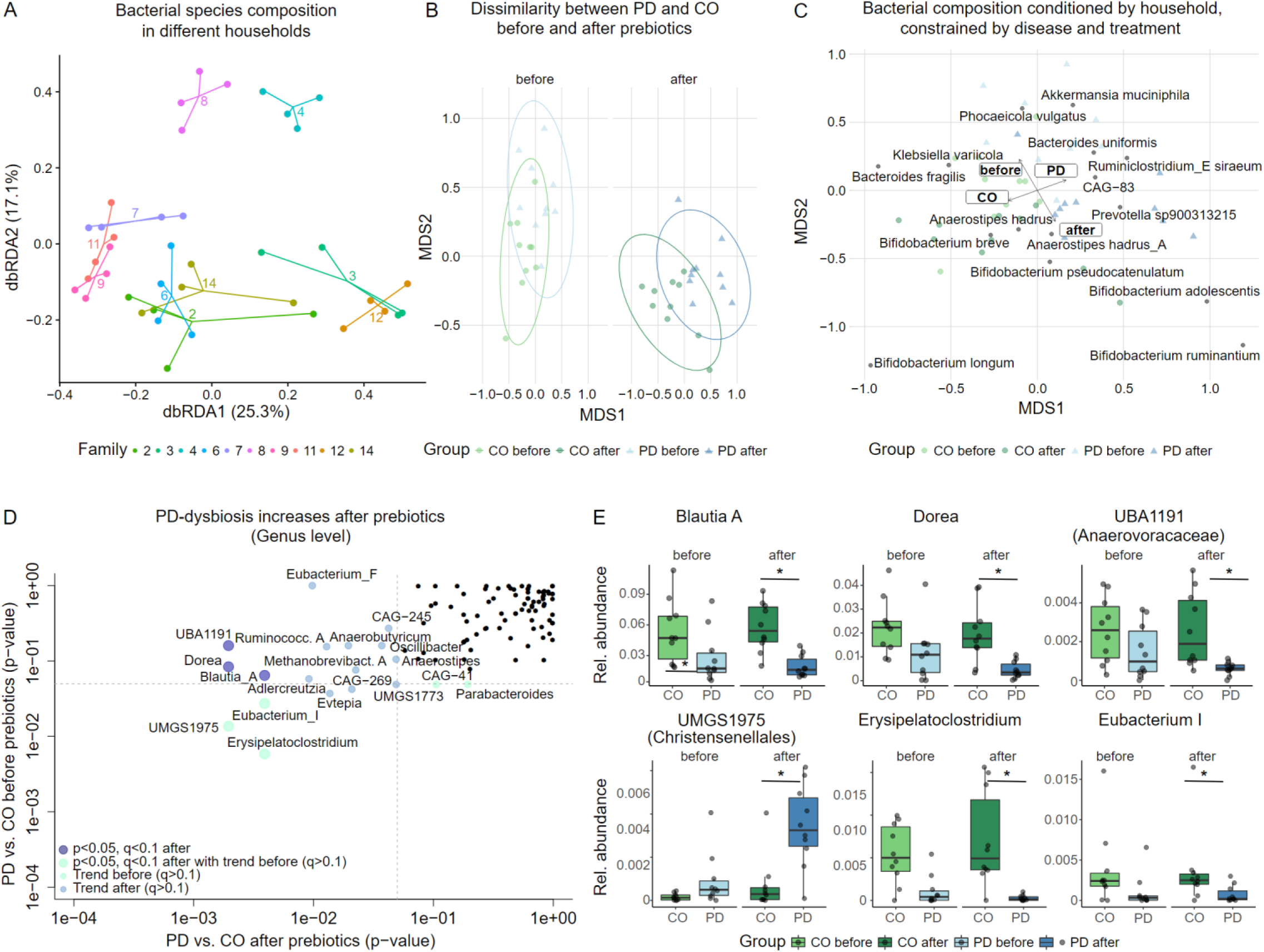
Household-effect and taxonomic differences between PD and CO. A, the dbRDA shows a strong clustering of metagenomes by household, i.e. from PD with their respective healthy spouse/CO, further termed “household-effect”; this effect explained 46% of data variability on MGS species level; B, dbRDA conditioned for households reveals a sign. difference in taxa dissimilarity between PD and CO after prebiotics, but not before prebiotics; C, top 15 correlated taxa (MGS species) between the constrained dbRDA (conditioned for households) and study group (PD/CO) or diet intervention (before/after prebiotics), indicating a strong association of different *Bifidobacteria spp.* with the intervention. D and E, several genera were markedly reduced in the PD group after prebiotics, paralleled by several trends, e.g., enrichment in *Eubacterium F* and *Methanobrevibacter_A* in PD; some of these Genera already tended to differ before prebiotics (p<0.05, but q> 0.1 before prebiotics; p<0.05, q<0.1 after prebiotics; data in D is presented with p-values generated by univariate tests between PD and CO on a log scaled axis, Wilcoxon signed-rank test; data in E is presented as relative abundance of the different Genera). * = p<0.05, q<0.1.

**Suppl. Table 2.**
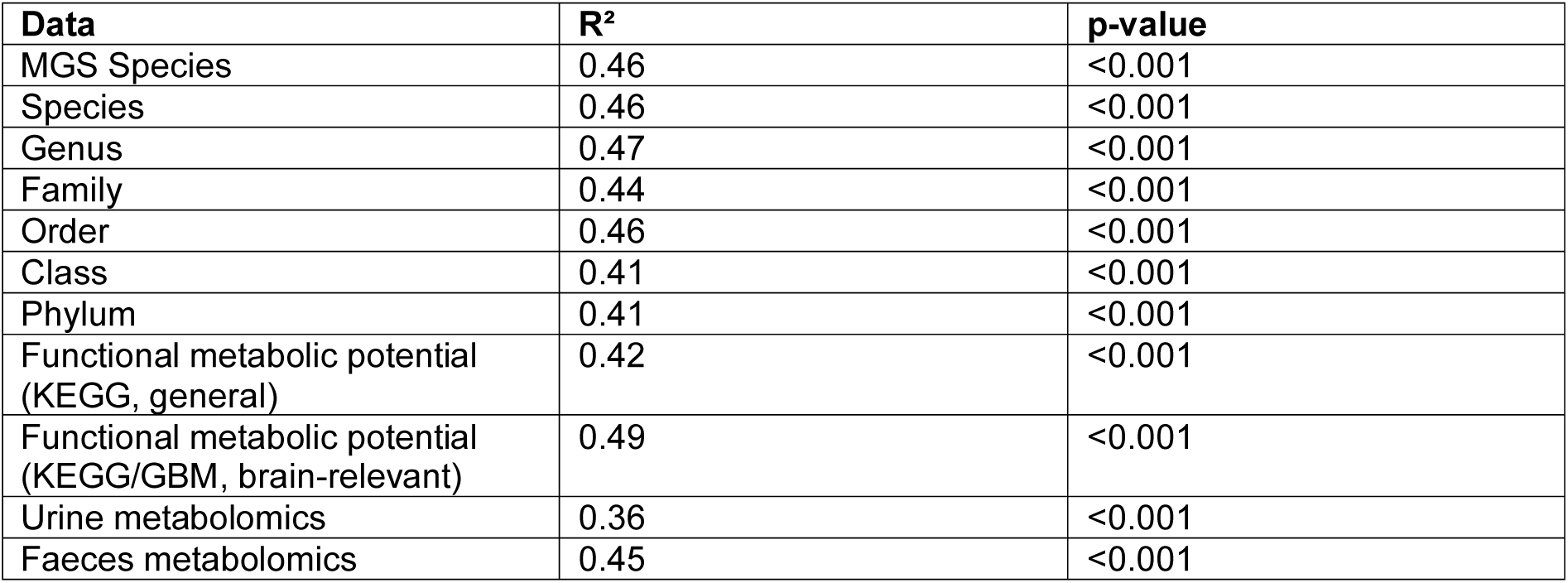
Household-effect on different data types assessed in this study.

| **Data** | **R<sup>2</sup>** | **p-value** |
| --- | --- | --- |
| MGS Species | 0.46 | <0.001 |
| Species | 0.46 | <0.001 |
| Genus | 0.47 | <0.001 |
| Family | 0.44 | <0.001 |
| Order | 0.46 | <0.001 |
| Class | 0.41 | <0.001 |
| Phylum | 0.41 | <0.001 |
| Functional metabolic potential (KEGG, general) | 0.42 | <0.001 |
| Functional metabolic potential (KEGG/GBM, brain-relevant) | 0.49 | <0.001 |
| Urine metabolomics | 0.36 | <0.001 |
| Faeces metabolomics | 0.45 | <0.001 |

For our further analysis we therefore consequently used statistics conditioned for household effect. An ordination conditioned by household revealed that the dissimilarity in taxonomic composition was driven by differences between PD and CO subjects (R²=0.27, p<0.001, permutation test), or differences between before and after prebiotics (R²=0.2, p<0.001, permutation test, Fig. 2b and c):

Several taxa were identified representing the previously documented PD dysbiosis, including *Akkermansia muciniphila* and *Prevotella spp*. (*sp900313215)* enriched in PD subjects, while *Bacteroides fragilis* and *Anaerostipes hadrus* were enriched in CO (Fig. 2c). The strongest signal relating to diet intervention was an enrichment of several *Bifidobacteria spp.* after prebiotics in both CO and PD (multivariate and univariate tests, Fig. 2c, Fig. 3a, and Suppl. Fig. 3c). In PD patients, we observed a significant enrichment in six different *Bifidobacteria spp.*, four of which were also increased in CO after prebiotics (Fig. 3a and Suppl. Fig. 3c Wilcoxon signed-rank test). Similarly, *UMGS1975* (Christensenellales) was enriched after prebiotics in both groups, but more so in PD subjects that were also enriched for *UMGS1975* before the intervention. *Streptococcus thermophilus* was reduced only in the PD group after the diet intervention.

**Figure 3.**
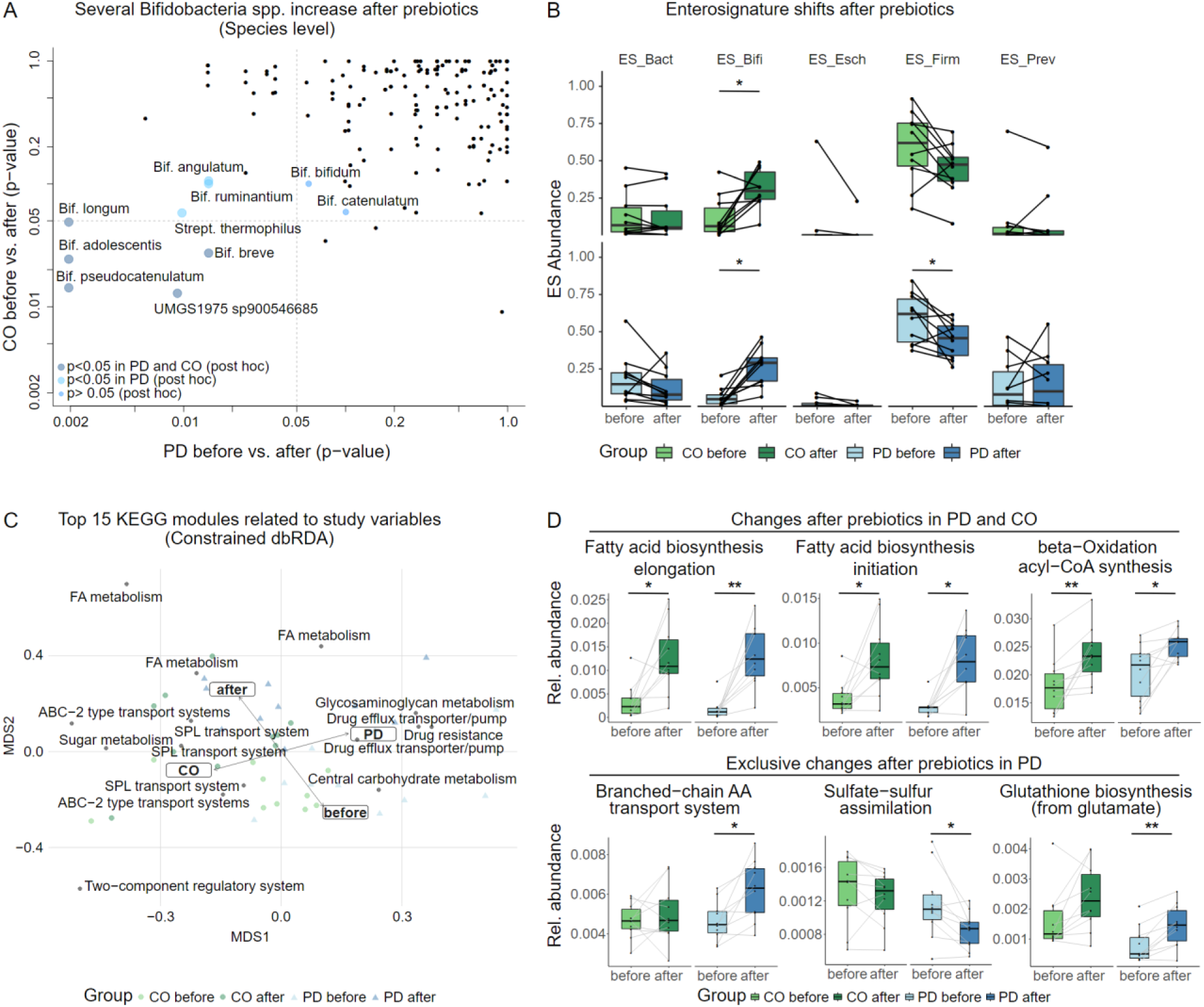
Gut bacterial communities affected by the prebiotic diet intervention. A, several taxa were significantly different between before and after prebiotics tests in both groups; n=6 *Bifidobacteria spp.* were enriched after prebiotics in PD, of which n=4 were also enriched in CO individuals; n=2 taxa are only changed in PD (Strept. Thermophilus, UMGS1975, data is presented as p-values generated with univariate tests/ Wilcoxon signed-rank test between before and after prebiotics on log scaled axes); B, ES composition shifted towards increased ES-Bifi in both groups while ES-Firmi decreased (sign. only in PD group, Wilcoxon rank sum test; C, top 15 KEGG modules in relation to the study group (PD/CO and diet intervention (before/after prebiotics), constrained dbRDA, conditioned for household),shows a strong correlation of fatty acid (FA) metabolism with the intervention (permutation test, R²=0.3, p<0.001) as well as several functions correlated to either PD or CO (permutation test, R²=0.16, p<0.001, SPL, Saccharide, Polyol and lipid transport System); D, Differences in KEGG Modules before vs. after prebiotics (relative abundance, Wilcoxon signed-rank test), increased microbial genes related to fatty acid metabolism after prebiotics were similarly observed in PD and CO subjects (upper panel), on the other hand several genes were only changed in PD (lower panel). * = p<0.05, ** = p<0.01 (AA, amino acid).

The PD-associated dysbiosis was not compensated by the diet intervention: *Blautia*, *Dorea, UBA1191* (Anaerovoracaceae) and *Erysipelatoclostridium* remained depleted in PD and *UMGS1975* (Christensenellales) remained increased in comparison to CO before and after prebiotics (all p<0.05, q> 0.1 before prebiotics; p<0.05, q<0.1 after prebiotics, Wilcoxon rank sum test, Fig. 2d and 2e). This was also reflected in a markedly different species composition between PD and CO after prebiotics (perMANOVA, R²=0.08, p=0.03 and R²=0.05, p=0.16, after and before, respectively, Fig. 2b). Notably, some subjects had elevated levels of genus *Klebsiella* (p>0.05), which decreased after prebiotics. Since this genus contains several pathobionts, this could be a benefit provided by prebiotics.

Alternatively, we cannot exclude that the high abundance of Klebsiella in some samples was due to inadequate anaerobic conditions during sample collection or storage.

To assess the extent of dysbiosis further, we assessed common bacterial guilds of the human gut microbiome (represented through Enterosignatures, ES) and their prevalence in our samples^27^. Overall, both study groups showed a good ES model fit, comparable between both groups (Suppl. Fig. 4a). In the PD group ES model fit increased marginally after prebiotics (p>0.05, Wilcoxon signed-rank test), while this increase was significant in CO (P=0.049, Wilcoxon signed-rank test, Suppl. Fig. 4a), indicating that the gut microbiome after the diet intervention is more similar to an average gut microbiome from the enterosignature model (trained on >5,000 gut microbiomes^27^).

**Suppl. Figure 4.**
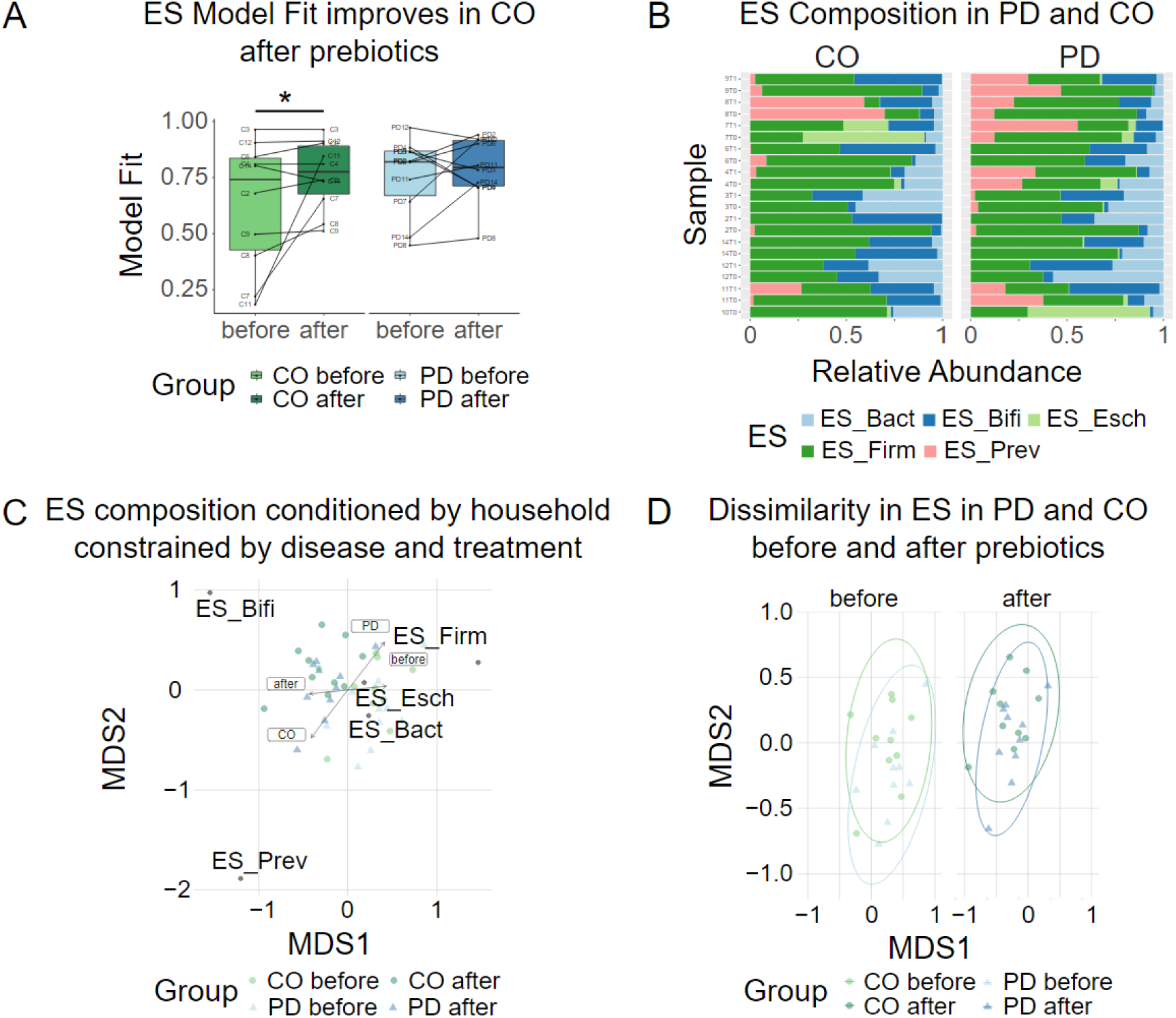
Enterosignatures. A, ES model fit is comparable between groups and most samples have a model fit above 0.75. Note that only the CO group showed an increased model fit after prebiotics (p=0.049,Wilcoxon signed-rank test); B, ES composition along study samples shown as the relative abundance of the respective ES per patient; C, Dissimilarity in ES composition was explained by the dietary intervention (R²=0.30, p<0.001, perMANOVA); D, Dissimilarity in ES composition showed no difference between PD and CO neither before nor after prebiotics (unconstrained dbRDA, Bray-Curtis distance). * = p<0.05

The dietary intervention had a strong impact on overall ES composition (Suppl. Fig. 4b), explaining 30% of variance amongst samples (after conditioning out the “household-effect”, R²=0.30, p<0.001, perMANOVA, Suppl. Fig. 4c). This was probably driven by an increase in ES-Bifi (Bifidobacterium dominated guild, p<=0.01 in both CO and PD, Wilcoxon signed-rank test), and decrease in ES-Firmi (Firmicutes dominated guild, p=0.08 and p=0.04 in CO and PD, respectively, Wilcoxon signed-rank test) after the prebiotic intervention (Fig. 3b). Note that this effect seemed overall stronger in PD than CO microbiomes and is potentially related to a higher intake of Lactulose in this group.

### The prebiotic intervention enhanced the bacterial metabolic potential for fatty acid metabolism in PD and CO and strengthened neuroprotective and antioxidative pathways in PD

Given the apparent consolidation of the PD dysbiosis following prebiotics, we sought to ascertain whether the intervention exerted any influence on the functional potential of the microbiome. For this we used key metabolic pathway modules relevant for gut bacterial metabolism from the KEGG database^28^, as well as GBM^29^ modules especially relevant for human brain functions. The functional composition of both, general and brain relevant pathways, was again dominated by household effects, which accounted for 42.1% and 49.1% of the total variance. In contrast, differences between PD and CO, explained 7.7% and 8.8% of variance in KEGG modules, while 7% and 6% of variance were explained through GBM modules, respectively (all p<0.001, perMANOVA). Interestingly, the dissimilarity for KEGG modules between PD and CO before prebiotics decreased after prebiotics, while GBM differences between PD and CO became more pronounced after prebiotics (perMANOVA conditioned for household, KEGG before: R²=0.10, p=0.028, after prebiotics: R²=0.08, p=0.048, GBM before: R²=0.07, p=0.025, after prebiotics: R²=0.10, p=0.003).

Investigating the modules driving these differences between PD and CO, we find the PD microbiome enriched in bacterial functions related to drug efflux and drug resistance, similar to our findings in 2017^6^. CO microbiomes were instead enriched in genes related to saccharide, polyol, and lipid transport systems (Fig. 3c). PD patients had increased abundances of genes related to butyrate synthesis, and decreased S-adenosylmethionine (SAM) synthesis (p<0.05, q>0.1, Wilcox rank sum test) compared to CO, but this was normalized after prebiotics (n.s. difference to CO, Suppl. material 1). After the prebiotic intervention, other functions differed between PD and CO, including Dihydroxyphenylacetic acid (DOPAC) Synthesis and GABAII Synthesis (decreased abundance in PD, p<0.05, q<0.1, Suppl. material 1) and Tryptophan degradation (increased abundance in PD, Wilcoxon rank sum text, p<0.05, q>0.1, Suppl. material 1).

We next determined bacterial functions changed with the prebiotic diet (before vs. after prebiotics comparisons). In both CO and PD, multiple fatty acid metabolism related genes increased in relative abundance after prebiotics, involved in the synthesis of medium- and long-chain fatty acids (Fig. 3c and 3d).

In addition, 19 KEGG modules related to nucleotide, amino acid, carbohydrate and lipid metabolism were significantly changed in both PD and CO after prebiotics (Suppl. material 1). Notably, the prebiotic diet decreased formaldehyde assimilation, which is identified as increased in the PD dysbiosis ^30^.

Compared to baseline values before prebiotics, several alterations in KEGG modules (N=20) were only different in PD patients (but not in CO) after prebiotics, e.g., Sulphate-sulphur-assimilation pathway associated genes decreased, genes related to glutathione (GSH), serine synthesis as well as branched chain amino acid transport increased (Wilcoxon signed-rank test, Suppl. material 1 and Fig. 3d), the latter matching our metabolomic observations (see below). Notably, several bacterial pathways leading to brain relevant metabolites changed after prebiotics (p<0.05, Wilcoxon signed-rank test, Fig. 4a and 4b). These could have potentially positive implications for PD patients, e.g. the changes in genes related to both the neurotoxic p-cresol and quinolinic acid (decreased synthesis and increased degradation, respectively). Inositol synthesis genes increased, and inositol degradation genes decreased (a neuroprotective metabolite).

**Figure 4.**
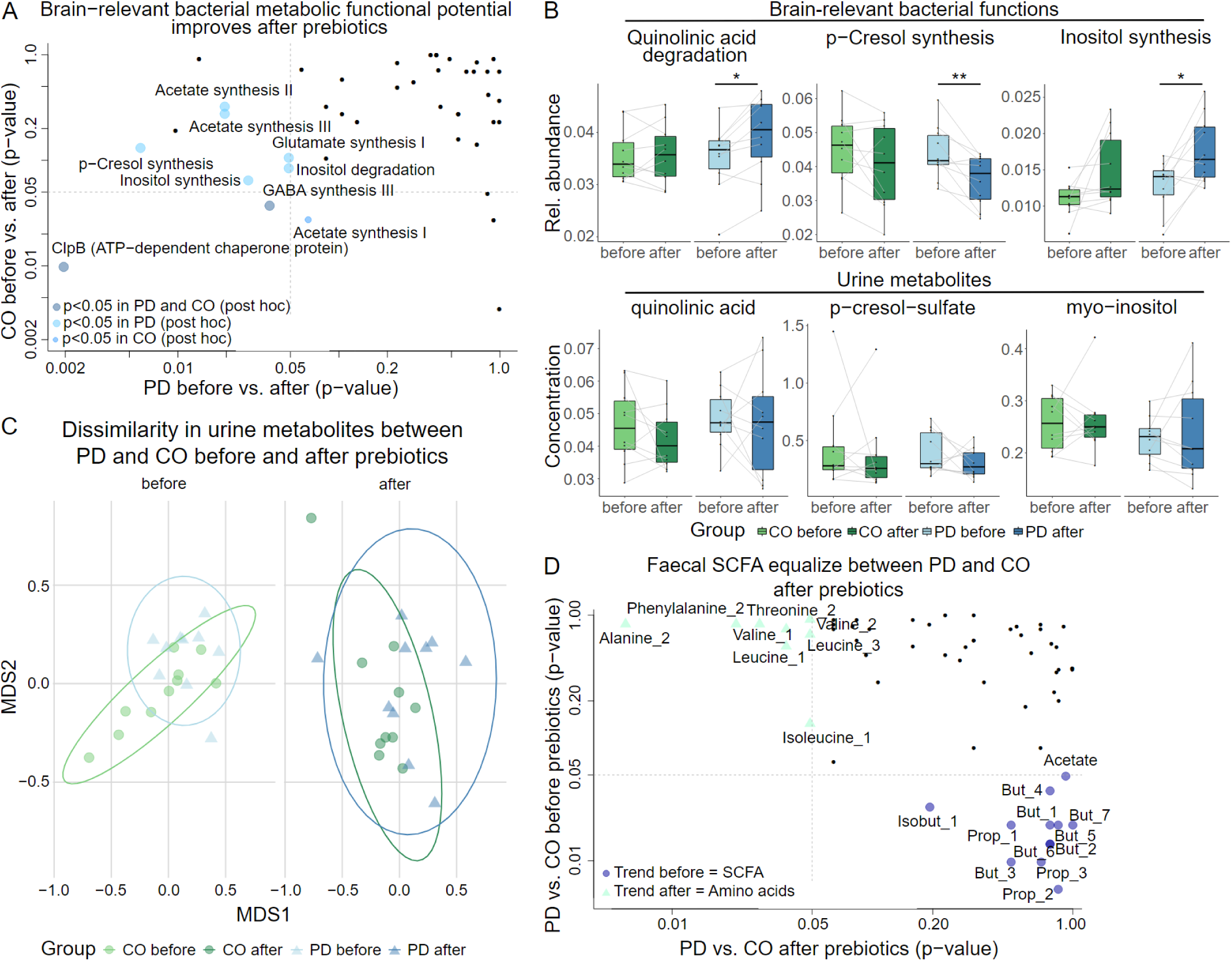
Functional metabolic potential and metabolite levels. A, brain-relevant functional metabolic potential (GBM modules) improves after prebiotics in PD in a potentially neuroprotective manner, e.g. p-cresol synthesis decreased and quinolinic acid degradation increased, both leading to neurotoxic compounds. X- and y-axis corresponds to p-values of Wilcoxon signed-rank test for changes due to the prebiotic intervention of CO (x-axis) and PD (y-axis) patients. B, the relative abundance of bacterial functional GBM modules improved only in PD patients after the prebiotic intervention compared to baseline before prebiotics (upper panel, p<0.05, q<0.1, Wilcoxon signed-rank test). This was also reflected in corresponding urinary metabolites (bottom panel, p>0.05, Wilcoxon signed-rank test). C, urine metabolite composition differed between PD and CO before prebiotics, but normalized after prebiotics (permutation test, before prebiotics: R²=0.08, p=0.044; after prebiotics: R²=0.05, p=0.36). * 0 p<0.05, ** = p<0.01; D, faecal metabolite profiles before prebiotics were different in SCFA’s between PD and CO. After prebiotics these differences are no longer observed, instead amino acid concentrations were different between PD and CO (all q >0.1, Wilcoxon rank sum test); But= butyrate, Isobut= isobutyrate, Prop= propionate.

### Bacteria driving changes in functional potential

To better understand the contribution of taxonomic changes to the observed changes in bacterial pathways, we correlated KEGG and GBM modules to MGS species clusters (partial correlation to correct for the time series). These functional metabolic changes could be correlated to different microbes, that were all previously implicated in PD^4,6^ (Suppl. material 2). Fatty acid synthesis (initiation and elongation) and beta-oxidation/acyl-CoA-synthesis correlated positively to several *Bifidobacteria* species, but negatively to *Alistipes spp*. and *Barnesiella spp*.. The former species contributes to the dysbiosis associated to PD, suggesting that an induced depletion of these taxa could benefit PD patients.

Several members of *Bifidobacteria* and *Oscillospiraceae* correlated to branched chain amino acid transport pathways (changed only in the PD group in before vs. after prebiotics comparisons), while e.g., *Blautia A spp.* showed a strong negative correlation. Further, *Eubacterium I spp*. were the strongest sign. positively correlating taxa with Sulphate-sulphur-assimilation (Suppl. material 2). Regarding brain-relevant GBM functions, a strong positive correlation was observed for different *Alistipes spp.* with inositol degradation as well as with p-cresol-synthesis, again suggesting that a diet-depleted *Alistipes spp.* would be of benefit in PD due to potentially reduced levels of neurotoxic compounds.

### PD faecal and urine metabolites profiles normalized with the diet intervention

Faecal metabolites^31^ approximate the bacterial metabolism in the gut, while urine metabolites^32^ can reflect both human and bacterial metabolism. We therefore analysed both faecal and urine metabolomes (NMR) of our cohort (N=80, coinciding with faecal sample collections). Variance in both urine and faecal metabolites was again mostly driven by differences between households (perMANOVA, R²=0.36, p<0.001, and R²=0.45, p<0.001; respectively, Suppl. Table 2), but study groups (PD vs. CO) had a sign. different urine metabolites composition (R²=0.04, p=0.03). Testing dissimilarities in urine metabolites for both timepoints separately (conditioning for households) revealed that the PD urinary metabolite profiles after the prebiotic intervention resembled the CO profiles (Fig. 4c). However, no clear causative biological signal was derived from the single metabolite composition to better explain this improvement based on differential abundance testing of all metabolites in urine between PD and CO. Comparing faecal metabolites between PD and CO before and after prebiotics revealed that several faecal SCFA normalized after prebiotics in the PD group (e.g., Iso-/Butyrate and Propionate); instead, amino acids concentrations differed, such as a decrease in Alanine and Iso-/Leucine (Wilcoxon Rank Sum test, p<0.05, q>0.1, Fig. 4d), or neurotoxic metabolites (but p>0.05, Fig. 4b), corresponding to the observed changes in the gut metabolic functional potential. Although limited by our cohort size, this highlights a plausible biological relation and potentially important result for PD patients.

## DISCUSSION

### Key results-Beneficial effects of the prebiotic diet for PD patients

The short-term prebiotic diet intervention was safe and well tolerated in all subjects (mild bloating was the only complain) and resulted in increased SCFA concentrations and improved gastro-intestinal symptoms in PD. We observed a trend towards reduced disease severity (UPDRSIII), which was inversely correlated with SCFA concentrations, indicating the importance of SCFA’s for gastro-intestinal functioning and possibly also motor symptoms in PD. Interestingly, in a recent study of faecal microbiota transplant^33^, the improvement in motor symptoms became pronounced between the sixth and twelfth month after the transplant, suggesting that the effects of a dietary intervention might also occur at a later point than assessed here.

We observed several gut microbes that were sign. different before and after the intervention between PD and CO metagenomes, and these coincided with several well-known “dysbiotic” PD gut microbes. However, on a functional and metabolic level, the prebiotic diet appeared to remove most of the aberrations observed between PD and CO patients, presumably improving gut microbial metabolism in PD patients: Apart from normalized SCFA concentrations in the faecal metabolite profile, we also found decreased branched chain amino acids (BCAA), potentially harmful for neurons by oxidative stress^34^, mitochondrial dysfunction^34^, and a decrease in dopamine synthesis^35^, while presumably neurotoxic compounds associated with PD^36,37^ were reduced in the urine metabolites after prebiotics (the aromatic amino acid p-cresol-sulfate, and the tryptophan/kynurenine downstream metabolite quinolinic acid). This coincided with changes in corresponding bacterial pathways derived from the metagenome. The prebiotic intervention seemed to restore a normal metabolite profile in general, as PD urinary metabolomic profiles were no longer distinct from CO after the prebiotic intervention.

Our analyses identified and had to be statistically corrected for a prevailing “household effect”. The importance of family shared microbes (possibly arising from social interactions, shared environments or diet) has been described before^38,39^, but the scale accounting for >36% (mean 44%) of explained variance at all data levels (Suppl. Table 2) was surprising to us.

### SCFAs at the Blood-Brain-Barrier

SCFAs, increased due to the prebiotic intervention in our cohort, have a range of presumed beneficial effects on gut- and blood-brain-barrier (BBB) integrity^9,40^. Translocating from the intestines to the systemic circulation, SCFA can reduce neuroinflammation^41^ and contribute to microglia maturation^42^ in murine models. Propionate, enriched in PD patients after prebiotics, has anti-inflammatory and BBB permeability-reducing effects in human brain endothelial cells^43^. Dietary fibre and SCFA can suppress intestinal inflammation, thereby improving the gut barrier function based on animal and cell culture studies ^44,45,46^. In addition to the observed changes in SCFA concentrations, we identified enriched bacterial pathways for fatty acid metabolism (middle-chain fatty acids, MCFAs, and long-chain fatty acids, LCFAs). MCFAs have been shown to have beneficial effects, particularly on metabolic features and insulin sensitivity ^47^, and LCFAs like palmitic acid (pathways enriched in our study), can lead to enhanced IgA antibody production *in vitro* and in mice^48^, contributing to host defence against pathogenic microorganisms.

### PD microbiome is not restored through prebiotics

To date, only few studies investigated the effects of dietary interventions on PD gut microbiome dysbiosis, focusing on constipation^49,50^ and/or pharmacokinetics^49^. The effects of dietary mixed fibres were investigated in a cohort^56^ of n=10 PD subjects over 10 days resulting in increased plasma SCFA, gut integrity markers and reduction in potentially pathogenic family (*Enterobacter*), while potentially SCFA-producing species (*Bifidobacterium*, *Faecalibacterium*) were increased, in accordance with our results after a 1-month intervention. With the inclusion of a control group, we could also characterize and monitor the PD-associated dysbiosis, corroborating our earlier work^4,6^.

The PD dysbiosis persisted during the diet intervention. Whether extended prebiotic use could restore the PD microbiome in the long-term, or if other, as-yet-unknown host-microbe or microbe-microbe mechanisms are responsible, remains a subject for further research. The microbiome is highly individual (or family) specific^38,39^ and distinct microbial communities might show divergent responses to prebiotics, indicating a hierarchical specificity towards gut microbes^51^, potentially limited by physico-chemical fibre characteristics. These mechanisms might be strain specific. Dysbiosis associated taxa (e.g., *Akkermansia*) in PD patients might stem from specific strains, which are particularly successful in outcompeting other microbiome taxa but with detrimental effects for the host. The removal of a persistent (dysbiosis associated) member of the microbiota might require much stronger remedies than a prebiotic, as most bacterial strains persist in the gut microbiome for extended periods^39^. Other factors that favour dysbiotic taxa in PD patients include constipation and increased transit time, as discussed in^4^. Although it is tempting to speculate, that the PD microbiota is less adaptable and remains therefore trapped in a dysfunctional state, we cannot exclude that the persistent dysbiosis might be a less favourable effect of the SCFA-enhancing diet in PD.

### Alterations in bacterial functional capacity with potential pathophysiological implications for PD

Following prebiotic intervention, alterations in gene abundance were identified in multiple bacterial metabolic pathways, particularly within the PD cohort. These findings offer a potential pathophysiological relevance to PD. This included a normalized SAM synthesis, enhanced GSH metabolism, decreased DOPAC synthesis, and an increased Tryptophan degradation along with the above-mentioned reductions in pathways resulting in neurotoxic metabolites (p-cresol sulphate and quinolinic acid) and increase in a pathway resulting in the neuroprotective metabolite myo-inositol. SAM, an endogenous amino acid metabolite involved in the methylation and trans-sulfuration processes critically modulates autophagy^52^, a process also relevant for PD, but excessive SAM on the other hand has been shown to cause PD like symptoms in rats^53^. It has been suggested that PD severity is related to a disturbed host-trans-sulphuration, a pathway central to antioxidant response, along with an increased bacterial Sulphur-metabolism^76^, related potentially to PD medication^54^. GSH (a SAM derived metabolite) is involved in neuro-immune and neuro-oxidative processes and in the regulation of cell death. Reduced GSH levels also is a key finding in PD brain tissue^55^, associated with mitochondrial dysfunction constituting a critical factor in the neuroinflammatory and -degenerative processes in PD^56^. The major dopamine metabolite, dihydroxyphenylacetic (DOPAC), seems to play a role in PD pathophysiology associated cell death^57^ and mitochondrial inhibition. Prebiotically decreased bacterial DOPAC concentrations in parallel with elevated antioxidative GSH concentrations in the gut lumen could in turn be advantageous for the human host with PD. However, it remains to be seen whether the aforementioned changes in bacterial metabolism or respective metabolite changes within the gut lumen will reach the human host in a manner that beneficially influences the PD process.

### The therapeutic potential for Bifidobacteria in PD

The prebiotic intervention led to a substantial increase in *Bifidobacteria.* Bifidobacteria were among the taxa consistently increased in PD patients^4,58^, albeit several studies^59,60^ controlled for laxatives such as the *Bifido*-genic Lactulose used here. These *Bifidobacteria* were most likely already colonizing the host’s gut and might therefore be more desirable for the PD patient, as seeding the gut microbiome with newly colonizing *Bifidobacteria* (or any other probiotic) could inadvertently negatively impact the gut microbiome, e.g. elicit an immune response to so-far-unknown taxa and/or disrupt the established and stabilized microbial ecosystem through the new strains. Prebiotics may represent a more suitable option for chronic conditions such as PD than probiotics, because of their longer-lasting effects^58,59^.

*Bifidobacteria* have the potential to benefit the PD patient and they are sometimes used as *Psychobiotics*^60^ (a subclass of probiotics), given their ability to stimulate neurotransmitters, SCFA, anti-inflammatory cytokines, production of γ-aminobutyric acid (GABA) or tryptophan ^61,62^ or enteroendocrine hormones with potentially brain-protective functions. However, *Bifidobacteria* could also be harmful, as their metabolism of tryptophan can produce potentially harmful metabolites (i.e., the neurotoxic quinolinic acid^63^) and certain *Bifidobacteria* strains can *in vitro* metabolize L-Dopa via a deamination^64^, potentially interfering with standard PD pharmacotherapy. Thus, despite many potential benefits that *Bifidobacteria* could provide, the actual species and strains already residing in a microbiome should be assessed before manipulating their abundances through prebiotic interventions.

### Limitations

Due to the limited number of subjects involved in this pilot study, the resulting data may be subject to bias and therefore require further corroboration. Lactulose was prescribed to PD patients as a basic medication to treat constipation at a dosage exceeding that used in the control group i.e., prebiotic dosage given for reasons of comparability. However, given our intention to investigate potential disease-modifying strategies that are particularly focused on PD patients, this approach may be deemed justifiable.

## CONCLUSION

A systematic investigation of metagenomics and metabolomics in both faecal and urine samples demonstrated that a SCFA-promoting diet modulated the metabolism of the intestinal microbiota in PD patients, along with improved gastro-intestinal symptoms. Hence, the potential of a microbiota-directed prebiotic intervention as a modifying therapeutic approach in PD appears promising. To safely exploit the therapeutic potential of *Bifidobacteria* in PD, we suggest using strain resolved metagenomics to guide prebiotic regimes to control for potentially harmful effects (such as L-Dopa deamination). It remains to be determined whether the dysbiosis associated with PD can be restored with longer-term prebiotic interventions or may require a probiotic approach or a (faecal) microbiome transfer. Larger placebo-controlled trials with longer-lasting dietary interventions including different types of fibres and probiotic interventions are required and justified given the beneficial metabolic changes observed in our data.

## METHODS

### Study design and participants

This prospective controlled clinical pilot study was approved by the local ethics committee of the University of Bonn, Germany, and all participants gave written informed consent (internal ethics vote 145/17).

The study was registered in the German Clinical Trials Register (DRKS under the number DRKS00034528). Inclusion criteria were: (1) idiopathic PD (mild to moderately advanced, i.e., Hoehn and Yahr 1-2) with (2) stable medication within the previous three months, and (3) willingness for their healthy non-PD spouses to participate; (4) both at ages ≤ 75 years. Exclusion criteria were (1) atypical parkinsonism; (2) PD patients with deep brain stimulation or continuous intestinal levodopa infusion; (3) chronic and inflammatory gastrointestinal diseases including severe chronic constipation; (4) the use of laxatives, antibiotics, or immunosuppressive agents in the past three months (note that n=1 PD took 1 day of antibiotics 4 weeks before study); (5) lactose intolerance; and (6) veganism. Participating PD patients were recruited from the Department of Neurology, University of Bonn, Germany, and underwent the dietary intervention together with their healthy spouses. The control group was selected to ensure a certain degree of standardisation and comparability with regard to the dietary recommendations provided throughout the course of the study. Furthermore, it was hypothesised that habitual diets would prove more comparable to those of age-matched healthy controls, thus enhancing the reliability of the baseline conditions. The recruitment of participants was conducted between October 2017 and July 2019, with the relevant study data collected concurrently. N=11 PD patients and their N=11 healthy spouses (control group, CO) were included in the study. One couple (1 PD, 1 CO) withdrew from the study before the final visit for reasons not related to the study.

PD was clinically diagnosed according to the UK Brain Bank criteria. Disease severity was measured using the Movement Disorders Society Unified Parkinson’s Disease Rating Scale (MDS_UPDRS) part III. Gastrointestinal symptoms and constipation presence were evaluated through a Gastrointestinal Symptom Rating Scale (GSRS ^65^, selected items: borborygmus, abdominal distension, increased flatus, decreased passage of stools, increased passage of stools, loose stools, hard stools, urgent need for defecation and feeling of incomplete evacuation, rated 0-3 based on intensity, frequency, duration, or social impact) and a post-prebiotic interview for adverse effects.

### Dietary intervention

All participants agreed to follow a recommended diet, a combination of nutrients rich in fibres, fruits, and vegetables for 4 weeks: a combination of i) 2 apples/10 apple rings a day (rich in pectin, a polysaccharidic prebiotic) and ii) 5 portions per week (portion= the size of the own hand) of foods rich in resistant starch, lignin, and anthocyanins (lentils, potatoes, green beans, onions, oat bran, lettuce, olive oil, and bananas as well as strawberries). PD patients were taking a prebiotic Lactulose syrup to treat constipation as a basic medication and the spouses thus were ingesting Lactulose in a prebiotic dosage. Lactulose, a non-digestible synthetic disaccharide, is used for different purposes, such as a prebiotic, for constipation and hepatic encephalopathy treatment (ranging from 10g to 100g respectively). PD patients took 2×15 mL (2×10 g) daily for treatment of constipation, while controls took 1×15 mL (1×10 g, i.e., prebiotic dosage). Lactulose is anaerobically fermented in the colon by microbiota, enhancing *Bifidobacteria*, *Lactobacillus* (and SCFA^66,67^). Dietary baseline information was assessed with a questionnaire (Suppl. material 3). Diet compliance was verified through self-maintained food logs over the 4 weeks, reviewed after completion of the study (n=7 representative diet days per subject, i.e., n=5 weekdays and n=2 weekend days across the 4 weeks). Diet diaries were analysed using the Nutritics dietary analysis software (Nutritics Ltd), based on the German Food Composition Database available within Nutritics. To establish a comparable and scalable measure for assessing dietary adherence, we focused on measuring the consumption of raw apples, as they are easily quantifiable, as well as total fibre intake. It should be noted, however, that apple/fibre consumption alone does not necessarily indicate full adherence to our recommendations. Due to the nature of our recommendation as a portion (resembling the size of the own hand) and because we did not systematically quantify dietary habits over a longer period before intervention, application of absolute values of other nutrients was considered inappropriate. The food score was calculated as following: the consumption of raw apples based on our given recommendations (2 Apples= 365 g/d or 10 Apple rings dried = 65 g/d) was considered 100% adherent. Then the fraction of the participants’ diet days with an apple consumption below the mean apple consumption of all participants was computed (mean=70%/d of our recommendation). A fraction of more than 0.75 days was considered as less adherent. Additionally, dietary analysis including estimating the average fibre intake of every representative day was calculated and used for basic statistical testing (see below), using the Nutritics dietary analysis software.

### Sample collection

Urinary and faecal samples were non-invasively collected at baseline and 4 weeks after diet. Faecal samples were put in a specimen collector (Sarstedt) and stored in a plastic bag under anaerobic conditions (Anaerocult® P, Merck Millipore). Participants were advised to collect samples the latest the evening prior to or at the day of the study visit. All samples were cooled in a refrigerator (5-7 °C) at the patients’ home and transported to the study site with a cool pack. Samples were immediately stored at - 80°C upon arrival at the study site. Additionally, routine laboratory parameters were assessed at baseline and 4 weeks after diet (focusing on serum sodium and potassium levels as safety measures due to the ingestion of Lactulose syrup).

### Outcome

Primary: i) modification of SCFA in faeces, ii) changes in gut microbial composition including key species for SCFA production, and iii) changes in bacterial metabolism based on bacterial genomes and metabolomics analysis in urine and faeces. Secondary: i) changes in motor symptoms (UPDRSIII), ii) modification of gastro-intestinal symptoms (modified GSRS, stool frequency, side effects).

### Faecal DNA Extraction and sample preparation

DNA extraction was performed with the Maxwell® RSC PureFood GMO and Authentication Kit (Cat. #AS1600) according to the manufacturer’s recommendations. In brief, 200 mg of faeces were placed into a 2 mL microcentrifuge tube and 1 mL of CTAB Buffer was added. Samples were heated at 95°C for 5 minutes and allowed to cool down for 2 minutes. Manually homogenisation was performed with bead beating in the 2 mL Lysing Matrix E tubes (containing 1.4 ceramic spheres, 0.1 silica spheres, and one 4 mm glass bead) using a Homogenizer (FastPrep, Setting/Speed 6.0, 3 times for 1.0 minute). Samples were then mixed with 40 μl of proteinase K and 20 μl of RNase A and incubated at 70°C for 10 minutes. Cartrides were prepared according to the manufacturer’s recommendations (300 µl lysis buffer). The Maxwell RSC was run with the PureFood Protocol, automatically purifying and eluting DNA in 100μl.

### Sequencing

Genomic DNA was normalised to 5 ng/µl with EB (10mM Tris-HCl). A miniaturised reaction was set up using the Illumina Nextera DNA Flex Library Prep Kit (Illumina Catalogue No 20018704). 0.5 µl Tagmentation Buffer 1 (TB1) was mixed with 0.5 µl Bead-Linked Transposomes (BLT) and 4.0 µl PCR grade water in a master mix and 5 μl added to a chilled 96 well plate. 2 µl of normalised DNA (10 ng total) was pipette mixed with the 5 µl of the tagmentation mix and heated to 55 ⁰C for 15 minutes in a PCR block. A PCR master mix was made up using 4 ul kapa2G buffer, 0.4 µl dNTP’s, 0.08 µl Polymerase and 4.52 µl PCR grade water, contained in the Kap2G Robust PCR kit (Sigma Catalogue No. KK5005) per sample and 9 µl added to each well need to be used in a 96-well plate. 2 µl of each P7 and P5 of Nextera XT Index Kit v2 index primers (Illumina Catalogue No. FC-131-2001 to 2004) were added to each well. Finally, the 7 µl of Tagmentation mix was added and mixed. The PCR was run with 72⁰C for 3 minutes, 95⁰C for 1 minute, 14 cycles of 95⁰C for 10s, 55⁰C for 20s and 72⁰C for 3 minutes. Following the PCR reaction, the libraries were quantified using the Quant-iT dsDNA Assay Kit, high sensitivity kit (Catalogue No. 10164582) and run on a FLUOstar Optima plate reader. Libraries were pooled following quantification in equal quantities. The final pool was double-SPRI size selected between 0.5 and 0.7X bead volumes using KAPA Pure Beads (Roche Catalogue No. 07983298001) and quantified on a Qubit 3.0 instrument and run on a D5000 ScreenTape (Agilent Catalogue No. 5067-5588 & 5067-5589) using the Agilent Tapestation 4200 to calculate the final library pool molarity.

Samples were sent to Novogene (Novogene (UK) Company Limited, 25 Cambridge Science Park, Milton Road, Cambridge, CB4 0FW, United Kingdom) to be run along with sample names and index combinations used. Faecal samples were shotgun sequenced (paired end) using an Illumina Novaseq 6000 at Novogene. Demultiplexed fastq’s were returned on a hard drive, and further analyzed with the Matafiler Pipeline^39^.

### Targeted SCFA measurements in faeces

Frozen aliquots of raw faeces were sent to an external laboratory (GANZIMMUN Diagnostics GmbH, Hans-Böckler-Str. 109, 55128 Mainz, Germany) to measure SCFA levels via GC-MS (GC; Perkin Elmer Clarus 680, MS: Perkin Elmer Clarus SQ8). In brief, 1 g of raw faeces was suspended and homogenized in 26% NaCl-solution (ACROS - 387640025). Then 900 mL 6M HCl were added for stabilization. 3 mL of the former were transferred into a plastic tube together with 30 µL of Istd-solution (100 µL 2-ethylbutyric acid in 9.9 mL 0.6 M HCl-solution). Samples were thoroughly mixed and 50 µl, placed into a Wheaton-Vial, and closed with a crimp-cap. Calibration standards of the respective SCFA were similarly prepared in 26% NaCl, stabilized with 6 M HCL, and treated analogously as the faecal samples. Then levels of SCFA were measured (Headspace-System: Perkin Elmer TurboMatrix 40, Column: Phenomenex Zebron ZB-FFAP, 30 m length, 0.25 mm ID, 0.25 μm width, gas: Helium, 80 kPa, injection-volume: 1 μL, temperature-program: 2 min at 45 °C, heating (15°C/min) to reach 250°C, heat 250°C for a duration of 10 min, Headspace-temperatures: Vial-Oven 60 °C, transfer line 110 °C, injection-pin 95 °C). Data was sent electronically with SCFA concentrations given in µmol per 1 g faeces.

### Non-targeted metabolomics analysis in urine and faeces

Nuclear magnetic resonance (NMR) spectroscopy was used for non-targeted metabolomics. A detailed description of the metabolomics method including urine sample preparation, NMR analysis and data processing is outlined in Heinzmann et al^32^. In brief, 150 µL of urine is mixed with 50 µL PO_4_-buffer (100% D_2_O) and 10 µL 4.5 M KF in D_2_O. To extract aqueous faecal metabolites, we homogenized 50 mg faeces in 1 mL H_2_O using ceramic beads (NucleoSpin, Macherey–Nagel, Dueren, Germany) and a TissueLyser (Qiagen, Hilden, Germany) mixing for 3 x 20 sec at 4500 rpm with a 10 sec cooling break (< 0°C). Subsequently, the homogenate was centrifuged (13.000 rpm for 10 mins at 4°C), the supernatant evaporated with a SpeedVac, the dried extract reconstituted in 150 µL H_2_O and mixed with 50 µL NMR buffer and 10 µL 4.5 M KF. Samples were immediately submitted to NMR analysis and the same workflow protocols were used for urine and faecal extracts^32^. For metabolite identification, we analyzed the quality control sample (i.e., a mixture of all samples in the study) as a representative sample of urine and faecal water extract respectively with a series of 2-dimensional NMR analyses as specified in^68^. Data were imported into Matlab software R2011b (Mathworks, Natick, MA, USA) and the water region removed, and spectra normalized^69^. Relative quantification of metabolites was done using the peak height of selected peaks, as identified by a peak picking algorithm^70^. As the urine metabolites were commonly characterized by multiple peaks, we chose the best representative peaks for data reduction prior to statistical testing.

### ELISA (faecal Calprotectin and Alpha-1-Antitrypsin

Quantitative determination of calprotectin and alpha1-antitrypsin in stool each was performed with an enzyme linked immunoassay from Immundiagnostik AG (Stubenwald-Allee 8a, 64625 Bensheim, Germany; calprotectin: IDK® Calprotectin ELISA, K 6927, K 6927.20, MRP (8/14, S100A8/A9); alpha1-antitrypsin: IDK® α1-antitrypsin ELISA, K 6760) following the manufacturer’s instructions.

### Genome reconstruction, taxonomic and functional profiling of metagenomes

MATAFILER ^39,71^ (https://github.com/hildebra/MATAF3) was used to process raw reads, assemble metagenomes, reconstruct metagenomics assembled genomes (MAGs) and dereplicate these to metagenomic species (MGS). Briefly, raw shotgun metagenomes were quality filtered using sdm v1.63 with default parameters^72^. Kraken2 ^73^ was used to remove human reads. Host-filtered metagenome reads, were assembled using MEGAHIT v 1.2.9^74^ and reads were backmapped onto the assembly using Bowtie2 v2.3.4.1^75^, genes predicted with Prodigal v2.6.1 with parameters ‘‘-p meta’’ and a gene catalogue clustered at 95% nt identity using MMseqs2^76^. Matrix operations on the gene catalog were carried out using rtk2^77^.

MAGs were calculated using SemiBin2^78^, their completeness and contamination estimated using checkM2^79^. Using a combination of SemiBin2 MAGs and canopy clusters^80^ (https://github.com/hildebra/canopy2), high-quality reference genomes (>80% completeness, <5% contamination) were dereplicated into metagenomic species (MGSs) using clusterMAGs (https://github.com/hildebra/clusterMAGs) in MATAFILER. Abundances of MGS in different samples were estimated based on conserved marker genes and their median abundances within the gene catalog.

### Enterosignature calculation

The reapplication of Enterosignatures (ES) were generated at https://enterosignatures.quadram.ac.uk/. The Genus abundance table was fitted to the five Enterosignatures model^27^ and taxa names were matched between the abundance table (H) and the five Enterosignatures W matrix, which gives the weight of each taxon in each signature. The W matrix and H have been modified so taxa names and orders in each match. ES are a set of five signatures that represent common microbial guilds complementary in their metabolism and found in all gut samples. The ES model allows to define a normal/homeostatic microbiome, i.e., a low model fit =< 0.4 could be suggestive of a microbiome in an atypical state (or at least in a state that was not well represented amongst the >5,000 gut microbiomes the model was originally learnt from). Brain related metabolic functional potential was assessed with a previously published database of manually curated gut-brain modules^81^ (GBMs, with each corresponding to a single neuroactive compound production or degradation process).

### Statistical analyses

Statistical analysis was conducted in R 4.1.3. Alpha-diversity and Beta-diversity indices and compositional analyses were calculated using the R-packages rtk^77^, phyloseq^82^, microbiome^83^, and vegan^84^, and data was visualized with ggplot2 and custom R scripts. For Alpha-diversity measures i.e., indices of diversity and evenness (Shannon, Simpson) and richness (Observed, Chao1, and ACE), sample count matrices were rarefied to 90% of the minimal sample sum to ensure even sampling depth. For Beta-diversity i.e., the quantification of sample dissimilarity (between PD and CO or between before and after prebiotics based on Bray-Curtis or Euclidean distance, see below) and taxonomic composition, taxa count matrices were normalized by dividing each feature by the respective total sample sum (TSS).

For univariate tests, features from the abundance matrix were removed that were present in less than four samples or had less than 0.001% relative abundance. Significance between PD and CO was tested with a paired samples Wilcoxon rank-sum (Wilcoxon signed-rank test, function “wilcox.test(…, paired=TRUE)”) test between couples for each time point separately (PD vs. CO before and PD vs. CO after prebiotics) followed by a multiple testing correction (Benjamini-Hochberg), to account for the “family effect”. For within-patient comparisons, a Wilcoxon signed-rank test (see above) was performed on all data (before prebiotics vs. after prebiotics), then multiple testing corrected (Benjamini-Hochberg). Post-hoc testing was performed using a Wilcoxon signed-rank test (PD before vs. PD after and CO before vs. CO after). All P-values were corrected for multiple testing using the Benjamini–Hochberg false discovery rate^85^ (referred to as q-value) using the function “p.adjust”. Differences in categorial metadata between PD and CO were tested with a Fisher’s exact test as implemented in base R (function “fisher.test”). For all univariate tests, a p-value <0.05 and a q-value <0.1 were considered statistically significant. Effect sizes were calculated using the R-package coin^86^ and rstatix, using the function “wilcox_effsize” and “coin::wilcoxsign_test”. The effect size r varies from 0 to close to 1, and we considered 0.10 - < 0.3 (small effect), 0.30 - < 0.5 (moderate effect) and ≥0.5 (large effect).

For estimation of dissimilarity between study variables we used (un)constrained and conditioned distance-based redundancy analysis (dbRDA, function “dbrda” in vegan^84^). As this function does not allow missing data values, we imputed NAs in metadata with the mean value of the respective variable (this was the case for n=3 GSRS and n=1 stool frequency values missing). Estimation of dissimilarity was based on Bray-Curtis distance for taxonomic and function tables (composition of TSS normalized data) or based on Euclidean distance for faecal SCFA, faecal and urine metabolomics on log-transformed data (log x+1).

A permANOVA (function “adonis2” in vegan) was used to test the significance of the constraints in the conditioned dbRDA distances. As the household effect had a major impact on data variability (see also results section), we blocked the perMANOVA test for the variable household (i.e., the pairs PD + healthy spouses) with the function “setBlocks” in permute^87^ and tested the dissimilarity either for the total data or the different timepoints separately. Dissimilarity matrices were then associated with different variables (study group, time point, metabolites, taxa) with the function “envfit” (with a permutation test as implemented in vegan).

For composition plots, taxa were summarized into a higher phylogenetic level (i.e., *Genus* level) and rare taxa were combined (with the function “microbiome::aggregate_taxa” or “microbiome::aggregate_rare”, respectively, as implemented in microbiome^83^/phyloseq^82^).

Partial correlation analyses between taxa and KEGG/GBM modules were conducted with ppcor^88^ with spearman correlation corrected for autocorrelation (with the function “pcor.test” the pairwise partial correlations between two variables (x, y) can be controlled for a third variable (z, the individual participant to avoid autocorrelation)) and the correlation matrix was visualized with corrplot^89^.

## Code availability

The underlying code for this study is available in GitHub and can be accessed via this link https://github.com/PDMicro/PDD-scripts.

## Data availability

The metagenomic sequencing data generated and/ analysed during the current study is available in the European Nucleotide Archive, ENA repository, PRJEB57228. Further data types are available upon request from the corresponding authors.

## Author contribution

**JRB** conceptualised and performed the study, acquired funding, wrote the analytical code and performed the formal analysis and drafted the manuscript. **SR** performed the calprotectin and alpha 1 antitrypsin ELISA, helped in formal analysis and critically revised the manuscript. **SSH** performed the NMR metabolomics in faeces and urine and helped in formal analysis of the respective data. **AD** helped in formal analysis of the Enterosginatures. **DN** helped in formal analysis of the household effect. **MT** and **DSL** formally analysed the food logs with the Nutritics software. **MCS** helped in interpreting the SCFA and nutrition data and critically revised the manuscript. **AN** funded the calprotectin and Alpha 1 Antitrypsin ELISA and critically revised the manuscript. **UW** helped in recruiting the study participants and in interpreting the data and critically revised the manuscript. **FH** performed the bioinformatic analyses of the sequencing data, supervised the statistical code writing, interpreted all data and wrote the manuscript with JRB. All authors read and approved the final manuscript.

## Acknowledgements

This study was funded by the Hilde-Ulrichs-Foundation for Parkinson’s research. This research was also supported by the BBSRC Institute Strategic Programme (ISP) Food Microbiome and Health BB/X011054/1 and its constituent project BBS/E/F/000PR13631, the BBSRC Core Capability Grant BB/CCG2260/1 and its constituent project BBS/E/QU/23NB0006, the ISP Decoding Biodiversity BBX011089/1, projects BBS/E/ER/230002A and BBS/E/ER/230002B, and Cellular Genomics Cellular Genomics BBX011070/1, project BBS/E/ER/230001A. The funders played no role in study design, data collection, analysis and interpretation of data, or the writing of this manuscript. JRB was supported by a local grant (FEMHabil) at the medical faculty of the University Hospital Bonn. FH was supported by European Research Council H2020 StG (erc-stg-948219, EPYC).

We thank Lia Mareen Taube for her support during the study performance.

## Competing interests

All authors declare no financial or non-financial competing interests.

## Notes

### Competing Interest Statement

The authors have declared no competing interest.

### Clinical Trial

DRKS00034528

### Author Declarations

The ethics committee of the University of Bonn, Germany, gave ethical approval for this work (internal ethics vote 145/17).

